# SARS-CoV-2 seroprevalence and detection fraction in Utah urban populations from a probability-based sample

**DOI:** 10.1101/2020.10.26.20219907

**Authors:** Matthew H. Samore, Adam Looney, Brian Orleans, Tom Greene, Nathan Seegert, Julio C Delgado, Angela Presson, Chong Zhang, Jian Ying, Yue Zhang, Jincheng Shen, Patricia Slev, Maclean Gaulin, Mu-Jeung Yang, Andrew T. Pavia, Stephen C. Alder

## Abstract

This project’s aim was to generate an unbiased estimate of the incidence of SARS-CoV-2 infection in four urban counties in Utah. A multi-stage sampling design was employed to randomly select community-representative participants 12 years and over. Between May 4 and June 30, 2020, surveys were completed and sera drawn from 8,108 individuals belonging to 5,125 households. A qualitative chemiluminescent microparticle immunoassay was used to detect the presence of IgG antibody to SARS-CoV-2. The overall prevalence of IgG antibody to SARS-CoV-2 was estimated at 0.8%. The estimated seroprevalence-to-case count ratio was 2.4, corresponding to a detection fraction of 42%. Only 0.2% of individuals who had a nasopharyngeal swab collected were reverse transcription polymerase chain reaction (RT-PCR) positive. The prevalence of antibodies to SARS-CoV-2 in Utah urban areas between May and June was low and the prevalence of positive RT-PCR even lower. The detection fraction for COVID-19 in Utah was comparatively high.

**Article Summary:** Probability-based sampling provides an effective method for robust estimates of community-based SARS-CoV-2 seroprevalence and detection fraction among urban populations in Utah.

## INTRODUCTION

By mid-October 2020, more than 38 million infections and 1 million deaths due to SARS-CoV-2 have been confirmed worldwide (1), but the real infection count is likely much higher and continues to be a point of significant uncertainty. Case reporting significantly underestimates the total number of SARS-CoV-2 infections, because of the under-detection of asymptomatic or mildly symptomatic individuals and variation in the use and availability of diagnostic testing. Serology provides an independent method to estimate the true cumulative incidence of SARS-CoV-2 infection, using immune response evidence as an indication of previous infection. Although seroprevalence has been touted as a more standardized way to estimate the incidence of SaRS-COV-2 infection across different populations, it also presents challenges because of inconsistencies in test performance and sampling methods.

In May 2020, we launched the Utah Health and Economic Recovery Outreach (HERO) project, in partnership with state governmental agencies, to collect community-based data on SARS-CoV-2 infection rates. Our goal was to estimate the cumulative incidence of SARS-CoV-2 infection to benchmark case detection in community populations based on public health surveillance. In addition to measuring SARS-CoV-2 seroprevalence, we collected nasopharyngeal swabs to concurrently estimate the prevalence of reverse transcription polymerase chain reaction (RT-PCR) positivity. We applied methods of recruitment and analysis to minimize bias and maximize relevance for policy-making. Herein we describe the results of the first phase of the project, which was conducted in the “Wasatch Front”, the major population center of Utah, comprising a chain of contiguous cities and towns stretched along the Wasatch Mountain Range.

## METHODS

### Sampling design and recruitment

The total estimated population of the four counties included in this serological survey – Utah, Salt Lake, Davis, and Summit – is approximately 2.2 million, representing about 68% of the entire state. Overall, 29% of the population is younger than 18, compared to 22% in the US as a whole (2). The fraction of residents of the 4-counties that are non-Hispanic white is 76%, which is higher than the US population of 60%. Between March 14th and June 30th, 2020, the four counties reported 17,316 cases of SARS-CoV-2 infection (3).

Participants were recruited and enrolled between May 4th and June 30th, 2020. The sampling frame consisted of a list of all residential addresses (N = 657,870) in the four counties curated by the state of Utah. The 657,870 total addresses were grouped hierarchically into 16,677 census blocks, 1,089 census block groups, 389 census tracts, and 229 groups of adjacent tracts (“tract groups”). We categorized tract groups into fifteen strata based on combinations of county, ethnicity, median age, and reported positive case count from the Utah Department of Health.

We used two address-based probability sampling designs that differed in intensity of recruitment and geographic clustering. Both methods followed a random sampling design. Our primary sampling design included 11,563 addresses that were selected by randomly choosing 26 of the tract groups from the 15 strata, weighted by tract group population. We then selected approximately 420 addresses from each tract group by first randomly choosing thirty census block groups per census tract group and then fourteen addresses per census block group. The geographic address clustering facilitated recruitment and data collection and followed methods recommended by the Centers for Disease Control and Prevention.

Our secondary sampling frame comprised 14,012 addresses. We selected these addresses by proportionately oversampling the same strata as our primary sampling frame and excluding the tract groups selected in our primary sampling frame. The secondary sampling frame allowed us to expand the pool of participants and to broaden the geographic reach within the four counties.

To recruit our sample, we sent each address a postcard and a letter encouraging household members to participate. Participants were asked to complete a household survey, and household members age 12 and older were invited to take an individual survey and to undergo testing for IgG antibody and viral RT-PCR at a specified mobile testing site. In our primary sampling frame, addresses were also visited at their home by a recruitment field team that attempted up to three in-person contacts. All household members who completed the survey and were tested received a $10 gift card.

Each mobile testing site location included four sequential drive-through stations. The first collected basic information about the individuals in the vehicle; the second conducted the viral RT-PCR via nasopharyngeal swab; the third conducted the IgG antibody via blood draw; and the last quality-checked participation, provided information about receiving test results, and responded to participant questions. The analyses described here are limited to individuals who completed the individual survey and underwent serological testing.

### Laboratory methods

Serum specimens were analyzed using the Abbott SARS-CoV-2 IgG assay performed on an Abbott Architect i2000 instrument (Abbott Laboratories), according to the manufacturer’s instructions. The SARS-CoV-2 IgG assay is a qualitative chemiluminescent microparticle immunoassay that detects IgG binding to an undisclosed epitope of the SARS-CoV-2 nucleocapsid protein. The assay relies on an assay-specific calibrator to report a ratio of specimen absorbance to calibrator absorbance. The assay can be interpreted as positive (ratio >1.4) or negative (ratio <1.4). The manufacturer reports a sensitivity of 86·4% (95% CI: 65.1, 97.1) after 7 days from symptom onset and 100% (95% CI: 95.9, 100) after 14 days, and a specificity of 99·6% (95% CI: 99.1 99.9) (4). This estimate of sensitivity was derived from 88 symptomatic patients. However, other studies have reported lower sensitivities using this assay, ranging from 85% to 97%, when used in the general population (5-7). We observed that 20 (83%) of 24 individuals who reported a prior positive SARS-COV-2 test more than 14 days prior to collecting the antibody test were seropositive. With the cut-off at 10 days, 25 of 30 (83%) participants were IgG antibody positive. Therefore, we assumed a sensitivity of 83% in our primary analysis.

SARS-CoV-2 viral RNA was detected in nasopharyngeal swabs using the cobas SARS-CoV-2 assay (Roche Diagnostics), following the manufacturer’s instructions. The cobas SARS-CoV-2 assay detects the SARS-CoV-2 nonstructural ORF1 a/b region unique to SARS-CoV-2 (limit of detection 1,800 copies/mL). ARUP Laboratories performed all testing at the University of Utah.

### Statistical methods

This surveillance project was designated by the University of Utah Institutional Review Board as non-research. Data analysis used a series of steps to account for the sampling design, nonresponse, demographic balance, and the sensitivity and specificity of the serology assay.

### Accounting for sampling design and non-response

We computed sampling design weights to account for varying probabilities of sampling of households (8). These weights depended primarily on the ratios of the numbers of sampled households to the total numbers of households within each stratum of the primary and secondary sampling designs. We computed three further sets of weights to account for nonresponse at the household, individual, and serology testing levels. Household response weights were determined from estimated propensities of household response based on characteristics of the census block group where the household was located. Individual response weights were determined from estimated propensities of response by individuals within households based on characteristics of the census block group and the primary household respondent. Serology response weights were determined from estimated propensities for the provision of a serology sample based on individual survey responses. Propensities were estimated separately in the primary and secondary sampling designs using nonparametric boosted regression for household and serology response and logistic regression for individual response (9). Estimated propensities for membership in the primary versus the secondary design were used to align the secondary sampling design’s characteristics to those of the primary sampling design. Multiplication of each of the weights described above provided two sets of comprehensive weights that accounted for the design and nonresponse for the primary and secondary sampling designs. The weights for two sampling designs were then scaled based on the proportions of respondents in the two designs to provide a single final set of weights for estimating seroprevalence across the 4-county area. To prevent extreme variation in weights, we truncated weights that were either less than 10% or more than 10-fold greater than the median weight. Finally, we used iterative proportional fitting to optimize agreement of the marginal distributions of age, sex, Hispanic ethnicity, and education level between the weighted study sample and the US census of the 4-county area (10).

### Data Analysis

The primary sampling units (PSUs) for data analysis were defined by 54 census tracts included in the primary sampling design and primarily by block groups in the secondary sampling design. For Summit County, sampling was performed without clustering at the household level in the secondary sampling frame, so the household served as the PSU. We modeled the relationship of seroprevalence to predictor variables (e.g., county, demographic and clinical factors, behaviors and attitudes) using survey weighted generalized linear models for binary outcomes with variability assessed based on replicate jackknife weights (11, 12). We tested for the presence of a detectable temporal trend in seroprevalence by including calendar time as a continuous variable in models relating seroprevalence to the Utah Department of Health case count and calendar time. These analyses showed no trend for an effect of calendar time. Hence, analyses for seroprevalence were performed without adjustment for calendar time.

We corrected estimates of seroprevalence for assay error by applying the formula: (P1 - (1-specificity))/(sensitivity + specificity - 1), where P1 is the estimated prevalence within a given category of a predictor variable provided by the generalized linear models. We then used the parametric bootstrap to account for the sampling error in the manufacturer’s estimate of specificity when presenting lower and upper 95% confidence limits for prevalence. We estimated the seroprevalence-to-case-count ratio by computing the ratio between the adjusted prevalence estimates described above to the weighted average case count rates corresponding to the respondent’s zip code 10 to 17 days prior to the respondent’s serology test as reported by the Utah Department of Health. We applied chi-square tests based on logit transformed estimated prevalences and their associated covariance matrix estimated by the parametric bootstrap to perform hypothesis tests comparing prevalence between categories.

## RESULTS

### Participant Characteristics

Between May 4^th^ and June 30^th^, 2020, 11,563 households were randomly selected for a combined mailed recruitment and home visit, and 14,012 households were randomly selected for mailed recruitment only. Altogether, 8,108 individuals from 5,125 households completed surveys and were tested for the SARS-CoV-2 antibody; of those, 5,791 individual participants were in the combined home visit and mailed recruitment frame and 2,317 were in the mailed recruitment only frame. See Tables 1 and 2 for characteristics of participating households and individuals. The median age of participants was 44 (interquartile range 30-62); only 9.3% of participants were age 12 to 18. Overall, 6.6% of participants self-reported ethnicity as Hispanic, compared to 15.3% of the four-county population based on census data. The source population also differed from participants with respect to age distribution and education level. Accounting for response bias through iterative proportional fitting resolved these differences in county-level marginal distributions (see statistical methods appendix).

**Table 1.** Characteristics of participating households and individuals, enumerated at household and individual levels.

| Household-level factors | Participating households<br>(No. = 5,125) | Participating individuals* (No. = 8,108) |
| --- | --- | --- |
|  | no. (%) | no. (%) |
| County |  |  |
| Davis | 1023 (20%) | 1703 (21.0%) |
| Salt Lake | 2695 (52.6%) | 4021 (49.6%) |
| Summit (Park City) | 283 (5.5%) | 345 (4.3%) |
| Utah | 1124 (21.9%) | 2039 (25.1%) |
| No. of household members who participated in project |  |  |
| 1 | 1738 (34.2%) | 1027 (12.7%) |
| 2 | 2277 (44.8%) | 3683 (45.4%) |
| 3 | 541 (10.6%) | 1307 (16.1%) |
| >=4 | 532 (10.5%) | 2091 (25.8%) |
| No. of household members less than 12 years of age |  |  |
| 0 | 3537 (70.3%) | 5407 (67.6%) |
| 1 | 589 (11.7%) | 1053 (13.2%) |
| 2 | 499 (9.9%) | 850 (10.6%) |
| 3 | 239 (4.7%) | 424 (5.3%) |
| >=4 | 169 (3.4%) | 269 (3.4%) |
| Primary language spoken in household |  |  |
| English | 4866 (96.3%) | 7785 (97.1%) |
| Spanish | 132 (2.6%) | 169 (2.1%) |
| Other | 55 (1.1%) | 61 (0.8%) |

\*Completed survey and tested for serum IgG
†Missing values: # of household members who participated in project=37, # of household members less than 12 years of age=92, Primary language spoken in the house=72.

**Table 2:**
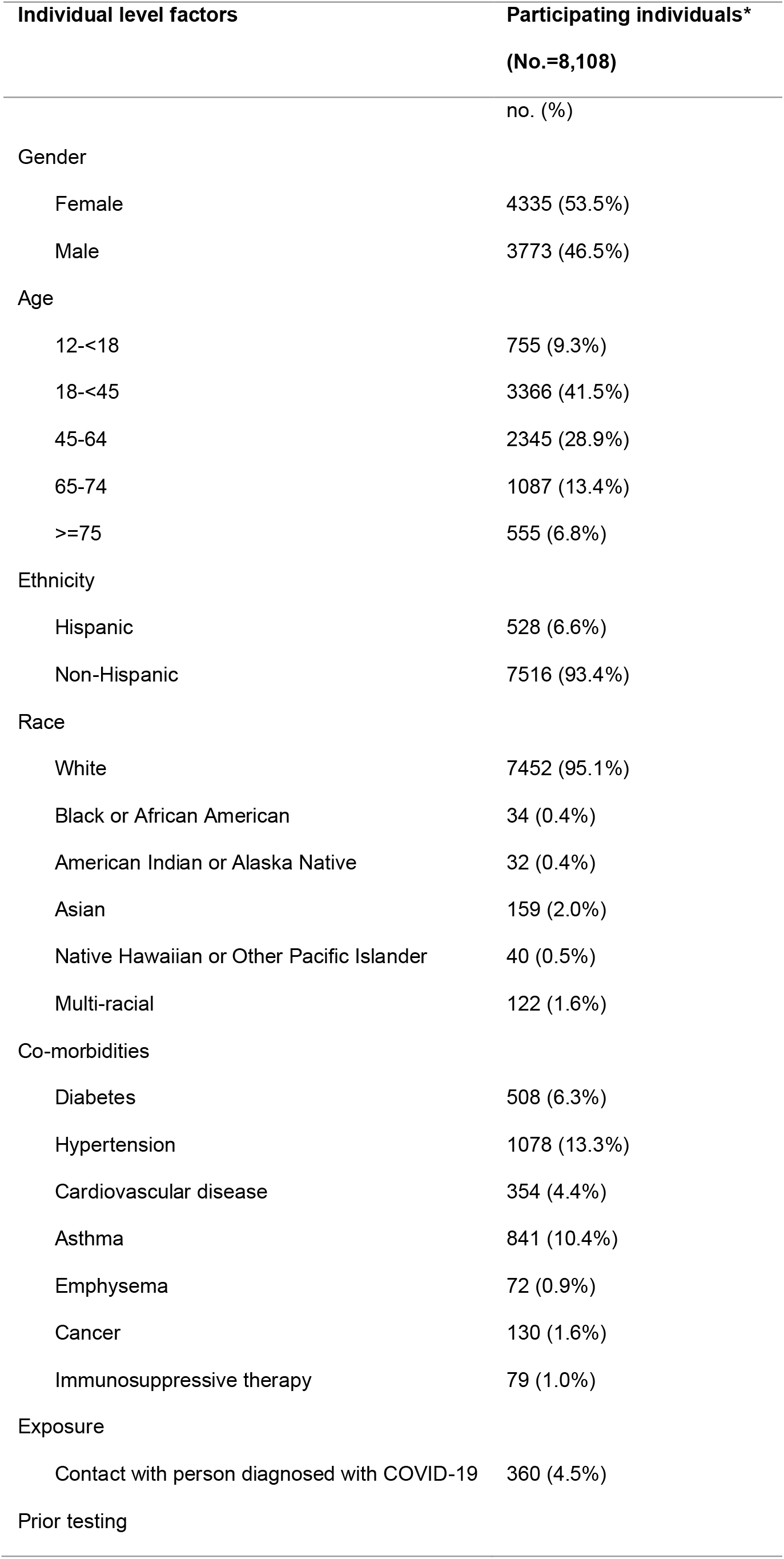
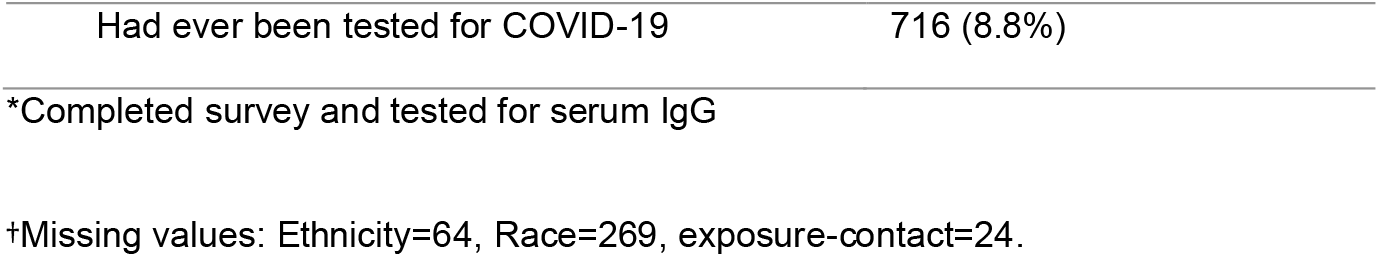
Characteristics of participating individuals*.

| Individual level factors | Participating individuals*<br>(No.=8,108) |
| --- | --- |
|  | no. (%) |
| Gender |  |
| Female | 4335 (53.5%) |
| Male | 3773 (46.5%) |
| Age |  |
| 12-<18 | 755 (9.3%) |
| 18-<45 | 3366 (41.5%) |
| 45-64 | 2345 (28.9%) |
| 65-74 | 1087 (13.4%) |
| >=75 | 555 (6.8%) |
| Ethnicity |  |
| Hispanic | 528 (6.6%) |
| Non-Hispanic | 7516 (93.4%) |
| Race |  |
| White | 7452 (95.1%) |
| Black or African American | 34 (0.4%) |
| American Indian or Alaska Native | 32 (0.4%) |
| Asian | 159 (2.0%) |
| Native Hawaiian or Other Pacific Islander | 40 (0.5%) |
| Multi-racial | 122 (1.6%) |
| Co-morbidities |  |
| Diabetes | 508 (6.3%) |
| Hypertension | 1078 (13.3%) |
| Cardiovascular disease | 354 (4.4%) |
| Asthma | 841 (10.4%) |
| Emphysema | 72 (0.9%) |
| Cancer | 130 (1.6%) |
| Immunosuppressive therapy | 79 (1.0%) |
| Exposure |  |
| Contact with person diagnosed with COVID-19 | 360 (4.5%) |
| Prior testing |  |
| Had ever been tested for COVID-19 | 716 (8.8%) |

\*Completed survey and tested for serum IgG
†Missing values: Ethnicity=64, Race=269, exposure-contact=24.

### Estimated seroprevalence

Eighty-nine individuals from 75 households were seropositive, corresponding to an unadjusted seroprevalence of 1.1% (Table 3). The four-county seroprevalence adjusted for sampling fraction, non-response, and test performance was 0.8% (95% confidence interval: (0.1-1.6)). Adjusted SARS-CoV-2 seroprevalence was estimated to be 5.7 % (95% CI 1.17-20.16) among individuals residing in households where the primary language was Spanish and 2.7% (95% CI 0.6-8.3) in individuals who self-reported as Hispanic; both estimates were significantly greater than the comparator groups. Seroprevalence was 4.45% in Summit County (which includes ski resort town Park City, an early infection hot spot in Utah), significantly higher than the other counties (p=0.03); the variation in seroprevalence across Utah, Salt Lake, and Davis counties was not statistically different.

**Table 3:** Overall and subgroup-specific seroprevalence.

| Demographic factors | Total<br>No. | Seropositive<br>individuals | Adjusted<br>seroprevalence* | Adjusted<br>seroprevalence |
| --- | --- | --- | --- | --- |
|  |  | no. (%) | % (95% confidence<br>interval) | P-value |
| **Overall** | 8108 | 89 (1.1%) | 0.8% (0.1-1.6) |  |
| **County** |  |  |  |  |
| Davis | 1703 | 16 (0.9%) | 0.1% (0-1.3) | 0.29 |
| Salt Lake | 4021 | 38 (0.9%) | 0.7% (0-1.8) |  |
| Summit (Park City) | 345 | 10 (2.9%) | 4.6% (1.0-15.1) |  |
| Utah | 2039 | 25 (1.2%) | 1.2% (0.1-3.4) |  |
| **Gender** |  |  |  |  |
| Male | 3773 | 41 (1.1%) | 0.7% (0-1.6) | 0.67 |
| Female | 4293 | 48 (1.1%) | 0.9% (0.2-1.9) |  |
| **Age** |  |  |  |  |
| Under 44 | 4119 | 39 (0.9%) | 0.9% (0.1-2.0) | 0.62 |
| 45 – 64 | 2345 | 31 (1.3%) | 0.8% (0.1-1.7) |  |
| Over 65 | 1642 | 19 (1.2%) | 0.4% (0-1.4) |  |
| **Ethnicity** |  |  |  |  |
| Non-Hispanic | 7516 | 75 (1%) | 0.5% (0-1.1) | 0.04 |
| Hispanic | 528 | 14 (2.7%) | 2.7% (0.6-8.0) |  |
| **Primary language spoken in**<br>**household** |  |  |  |  |
| English | 7785 | 78 (1%) | 0.5% (0-1.2) | 0.02 |
| Spanish | 169 | 11 (6.5%) | 5.7% (1.2-19.4) |  |
| **No. of household members**<br>**who participated in project** |  |  |  |  |
| 1 | 1027 | 15 (1.5%) | 0.7% (0-1.8) | 0.66 |
| 2 | 3683 | 35 (1%) | 0.5% (0-1.7) |  |
| >=3 | 3398 | 39 (1.1%) | 1.0% (0.2-2.3) |  |
| **No. of household members**<br>**less than 12 years of age** |  |  |  |  |
| 0 | 5407 | 64 (1.2%) | 0.6% (0-1.3) | 0.11 |
| 1 | 1053 | 11 (1%) | 1.1% (0-6.5) |  |
| 2 | 850 | 3 (0.4%) | 0.32% (0-3.0) |  |
| >=3 | 693 | 6 (0.9%) | 1.7% (0-13.7) |  |
| **Cumulative incidence per 1,000** |  |  |  |  |
| **in participant's zip code** |  |  |  |  |
| ≤ 200 per 100,000 | 3718 | 26(0.7%) | 0.2% (0-0.9) | 0.02 |
| 200–500 per 100,000 | 3012 | 34(1.1%) | 0.8% (0.1-2.0) |  |
| > 500 per 100,000 | 1378 | 29(2.1%) | 2.2% (0.6-5.5) |  |

\*Adjusted for sampling design and test sensitivity (0.83) and specificity (0.996)

Seroprevalence correlated with cumulative incidence estimated on the basis of reported case counts (Table 3). The adjusted seroprevalence was 2.1% in zip codes where cumulative incidence calculated from reported cases was greater than 500 per 100,000 population compared to 0.7% in zip codes in where the reported cumulative incidence was less than or equal to 200 per 100,000 population. The overall seroprevalence-to-case count ratio was estimated to be 2.4 (95% confidence interval 0.3-5.1), corresponding to a detected fraction of 0.42. This ratio was not statistically different across the four counties.

### Other descriptive analyses

Contact with a person who was diagnosed to have COVID-19 disease was reported by 360 (4.4%) participants, of whom 26 (7.2%) were seropositive (Table 4). Fourteen percent of individuals who reported contact with a family member with known SARS-CoV-2 infection were seropositive; in contrast, none of 38 individuals who reported exposure to SARS-CoV-2infection in their role as healthcare worker were seropositive. An analysis of 62 households with at least two members tested revealed 53 households with exactly one seropositive member and nine households with greater than one seropositive member. Twenty-three (19%) of the 123 members of these 62 households were seropositive, an estimate of the probability of infection given that another member of the household is infected. Assuming that infection for one of the infected members of each household was imported and that other household cases were transmissions from the index member of the household, the secondary attack rate was estimated to be 12%.

**Table 4:** Relationship between exposures and serological results.

| Exposures | Total<br>No. | Seronegative<br>individuals<br>Total No.=8019 | Seropositive<br>individuals<br>Total No.=89 | Adjusted<br>seroprevalence* |
| --- | --- | --- | --- | --- |
|  |  | no. (row %) | no. (row %) | % (95% confidence interval) |
| Contact with person diagnosed with COVID-19 | 360 | 334 (92.8%) | 26 (7.2%) | 8.5% (3.3-19.5) |
| Participant's relationship with contact |  |  |  |  |
| Family member | 97 | 83 (85.6%) | 14 (14.4%) | 14.8% (4.0-40.8) |
| Friend | 42 | 38 (90.5%) | 4 (9.5%) | 14.0% |
| Healthcare worker | 38 | 38 (100%) | 0 (0%) | 0.0% |
| Co-worker | 105 | 102 (97.1%) | 3 (2.9%) | 3.4% |
| Other | 78 | 73 (93.6%) | 5 (6.4%) | 3.1% (0.3-12.9) |
| Reside in household with at least one other member who is seropositive | 123 | 100 (81.3%) | 23 (18.7%) | 24.9% (10.5-48.7) |

\*Adjusted for sampling design and test sensitivity (0.83) and specificity (0.996). Confidence intervals are omitted for subgroups with fewer than 5 seropositive individuals.

Overall, 798 (9.9%) individuals reported having a prior test for coronavirus. Of 30 individuals who reported having had a positive SARS-CoV-2 test 14 or more days prior to collection of the serology test, 25 (83%) were seropositive, the figure that was used to estimate the sensitivity of the serological assay. Twenty-eight percent of seropositive individuals reported a prior positive SARS-CoV-2 test. If we assume a true seroprevalence of 0.8%, and assay sensitivity and specificity of 83% and 99.6%, respectively, the corrected point estimate for the detection fraction based on history of a prior positive RT-PCR test is 0.28/0.614 = 0.46, a value which is close to the estimate of detection fraction based on the seroprevalence-to-case count ratio.

Overall, 14 of 6251 (0.2%) individuals from whom a nasopharyngeal swab was collected had SARS-CoV-2 virus detected by RT-PCR; nine (64.3%) of these individuals were seropositive. The small number of positive RT-PCR tests precluded statistical analysis of factors associated with positivity or adjustment for response bias.

## DISCUSSION

Using a statistical sampling frame and adjusting for test performance and non-response, we estimated the prevalence of IgG antibody to SARS-CoV-2 in four urban counties in Utah between May and June to be only 0.8%. Thus, consistent with other community surveys, the large majority of the population lacked immunity to SARS-CoV-2 infection. Comparing seroprevalence to the cumulative incidence of SARS-CoV-2 infection based on case reporting, the estimated ratio of total to detected cases was 2.4, corresponding to a detection fraction of 0.42. We found higher seroprevalence in Summit County (4.5%), which is compatible with the extensive outbreak in the resort community of Park City that began in March. Seroprevalence was higher among persons who identified as Hispanic than non-Hispanic (2.7% vs 0.4%), and even more elevated in persons who lived in a household where Spanish was the primary language (5.7% vs 0.5%). This finding adds to the substantial body of evidence regarding ethnic and racial disparities in the spread of SARS-CoV-2 across populations.

Our estimates of seroprevalence and of the seroprevalence-to-case count ratio are generally lower than what has been reported in Utah and elsewhere in the US during this time period. A number of seroprevalence studies conducted in the US and other countries have been published to date (13-23), using a variety of assays and sampling methods (24). Some have relied on convenience samples or did not adequately control for response bias. The specificity of serologic methods for SARS-CoV-2 varies widely, and in a low prevalence population this can lead to substantial overestimation.(25) Not all studies have adjusted for test performance. These differences in methodology makes comparisons between studies challenging.

Our project, which involved random sampling of more than 25,000 households and use of intensive recruiting methods, is one of the largest to date. Our analytical approach accounted for multiple sources of error, including response bias and imperfect test performance. We were also able to generate an internal estimate of the detection fraction using self-reported histories of prior RT-PCR test results. After accounting for test error, the estimate of the detection fraction based on individual histories was 0.46, a value that corroborates our population estimate of the detected fraction (0.42).

We used a serological test that is reported by the manufacturer to have a specificity at 99.6%; however, even at this level of accuracy, statistically accounting for false positives is necessary given the low population prevalence of IgG antibody to SARS-CoV-2. To better account for the possibility of reduced sensitivity when asymptomatic infections are included, (26),we assumed a sensitivity of 83%, based on an analysis of project participants who reported having had a positive RT-PCR test in the past. Another factor that may limit sensitivity of the serum IgG to detect cumulative infection is waning immunity, which may be more prominent in those with mild or asymptomatic infection (26). In our study, serum was collected within two months following the previous RT-PCR SARS-CoV-2 test in 83% of individuals who reported having had a prior test.

With these considerations in mind, our estimate of the detection fraction is substantially higher than what has been reported in other serological surveys. A study that used residual clinical samples to measure SARS-CoV-2 antibody in 10 sites in the US estimated a detection fraction of 0.10 for residents of the US (16). That study estimated the seroprevalence in Utah at 2.2% with confidence intervals (1.2-3.4) that overlap our estimate. Similarly, our estimate of seroprevalence is lower than what has been reported in most other geographic regions. In a recent meta-analysis that reviewed 14 studies, only one region, southern Brazil, had an adjusted seroprevalence that was less than 1% (27). In a recently reported study, the projected prevalence of SARS-CoV-2 antibody in the US adult population was 9.2%, based on an analysis of 28,000 dialysis patients; in Utah it was 3.1%. Discrepancies between results of other studies and our findings are likely due in part to our sampling frame and recruitment methods and statistical methods that minimize bias (28). In our sample, seroprevalence is associated with increased work and activity outside the home and among individuals of lower socioeconomic status. In using an address-based sample, stratification of sampling based on demographic characteristics of the population, and intensive efforts to recruit participants, our sample better reflects the population than convenience-based samples. However, our results also suggest that Utah’s public health response to SARS-COV-2 was effective in case detection. Factors that likely contributed to the success of Utah’s approach to case detection include early expansion of access to testing, mobile testing that targeted heavily impacted communities, and a strong commitment to contact tracing and contact testing by the state and local health departments. This conclusion is also supported by our finding that 29% of seropositive individuals reported exposure to a known case.

The results of our analyses of clustering of seropositivity by household and of self-reported contact history are broadly consistent with each other. Our estimate of the secondary attack rate in households (12%) is similar to the prevalence of seropositivity among individuals who self-reported contact with a household member diagnosed with SARS-COV-2 and comparable to household secondary attack rates reported in other studies (29).

We observed that seropositivity was much more frequent than RT-PCR positivity, a finding that contrasts with selected other studies that combined viral detection and measurement of seroprevalence. For example, among randomly sampled residents of Indiana, the unadjusted prevalence of a positive RT-PCR was 1.74% compared to an unadjusted seroprevalence of 1.01%. The ratio of prevalence of antibody detection to prevalence of viral detection as observed in our community survey suggests that infections were accumulating linearly rather than exponentially during the period of study.

Several limitations are important to acknowledge. This paper covers the early period of the COVID-19 pandemic, reflecting the cumulative incidence of SARS-CoV-2 infection through mid-June. An updated analysis is needed to examine the secular trend in seroprevalence and determine whether the detection fraction continues to be high. Additional data will also enhance the feasibility of examining hot spots that may be geographically localized. Our analysis is not able to fully account for all sources of bias, particularly those factors that influenced the decision to participate at the household level.

In summary, we employed a project design where i) all participants were randomly selected; ii) antibodies were detected with a highly specific assay; iii) rigorous analytical methods were applied to account for bias and test error; and iv) population-level inferences were supported by analysis of survey responses. The most distinctive finding in our analysis was that the estimated total-to-reported case ratio was only 2.4, corresponding to a detection fraction of 42%. Further analysis is needed to determine whether this pattern has continued to hold up in subsequent months and to further assess the factors that influence SARS-CoV-2 transmission and detection.

## Data Availability

Individual level data will not be publicly available but aggregated data will be.

## Appendix--Statistical Methods

### Accounting for the complex survey design and assay error

Our statistical analyses incorporated a number of steps to account for nonresponse, demographic balance, and the sensitivity and specificity of the serology assay. We describe these steps below.

#### Step 1) Accounting for the sampling design

We estimated the probability that a household was sampled in the primary sampling design as

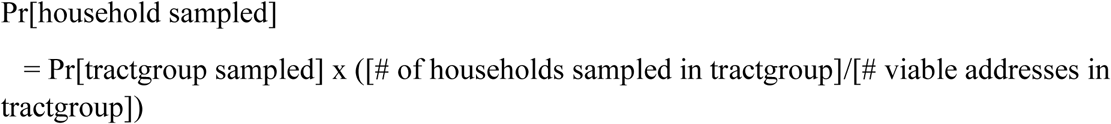

In strata for which more than one tractgroup was sampled, we approximated the probability that a given tractgroup was sampled as the product of the number of tractgroups sampled in that stratum and the probability of selection on a single draw. In the secondary sampling design we approximated the probability that a household was targeted for sampling as the proportion of viable households within each stratum that were designated for sampling.

#### Step 2) Accounting for Nonresponse

We estimated probabilities of response based on propensity models which used available information at the household, individual, and serology testing levels. The propensity models were fit separately for the primary and secondary sampling designs using the predictor variables listed in Table S1. We used boosted regression as implemented in the R TWANG statistical package (9) to estimate the propensities for a sampled household to respond to the household survey and for an individual survey respondent to provide serology samples. We used logistic regression to estimate the propensities for individuals to provide individual survey results among responding households. We computed weights to adjusted for overall nonresponse to serology testing as:

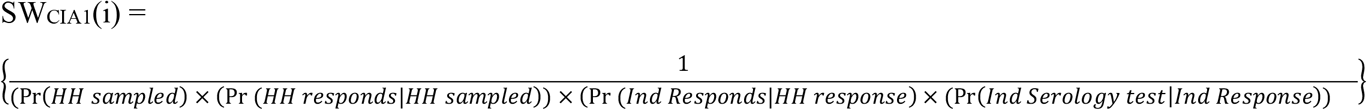

where Pr(*HH sampled*) represents the sampling design probabilities for each household, and where Pr(*HH responds*|*HH sampled*), (Pr (*Ind Responds*|*HH response*), and Pr(*Ind Serology test*|*Ind Response*) represent the propensity scores for household, individual, and serology response, respectively (8).

#### Step 3) Aligning Secondary Sampling Design to the Primary Sampling Design

The primary sampling design included both mail-push-to-web survey and in-person interviews, providing a duplicative contact strategy with two modes of contact, whereas the secondary sampling design includes only the mail-push-to-web survey. Thus we considered the primary sampling design to be less susceptible to non-response bias than the secondary sampling design. Therefore, we estimated a further set of propensity scores to reweight the individuals providing serology samples in the secondary sampling frame to align the characteristics of the of the secondary sampling design to the primary sampling design. These propensity scores defining these weights were also estimating using boosted regression, using the following predictor variables obtained from responses to the individual survey: 1) respondent’s sex, 2) respondents age, 3) respondent’s Hispanic ethnicity, 4) respondent’s education, 5) believes social distancing is important, 6) works outside the home at least a few times per week, 7) level of Covid concern 8) self-reported general health 9) self-report of being sick since March 1, 2020, and 10) known contact with someone who was diagnosed with COVID-19. After obtaining propensity scores, we computed 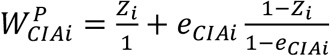 for each individual who provided a serology sample, where *Z*_*i*_ indicates membership in the primary Sampling Design. We then updated the sampling weights as:

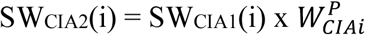

#### Step 4) Averaging weights across sampling designs

We treated the weighted samples from the primary and secondary sampling designs as both representing the same population. Accordingly, we computed the weighted average of the weights across the primary and secondary designs based on the proportion of respondents from each individual Sampling Design relative to the total number of respondents.

#### Step 5) Weight trimming

We implemented weight trimming to reduce the variability in the sampling weights separately in each county (12). Weights that were less than 10% of the median weight were increased to 10% of the median, and weights that exceeded the median weight by a factor of more than 10 were reduced to 10 times the median.

#### Step 6) Iterative Proportional Fitting

Because nonresponse adjustments are limited to variables known at each step, imbalances in known characteristics may still differ between the sample and target population even after applying the nonresponse weighs. Hence, we applied an additional calibration step by implementing iterative proportional fitting (often referred to as raking) to align the marginal distributions of age, sex, Hispanic ethnicity, and education between the weights study sample and the population of the 4 county target population (10). The population marginal distributions were derived using the 2018 Census American Community Survey 5-year estimates. The raking step was implemented using the following categorizations:

- Age, categorized as 12-29, 30-59, 60+, by county
- Sex, categorized as male and female, by county
- Ethnicity, categorized as Hispanic vs. Non-Hispanic, by county but excluding Davis County due to insufficient sample size.
- Education, categorized as completing 4-year college vs. all others (including those less than 25), by County.

#### Definition of Strata and Primary Sampling Units

In addition to incorporating the appropriate weights, statistical analyses must also account for the strata within each sampling design, as well as clustering of outcomes between different individuals in the same primary sampling units (PSUs) within the same stratum. The information on the amount of variation in seroprevalence between the census tract groups, the true PSUs of the primary sampling design, was limited, as the primary sampling design had only 26 census tract groups across 15 strata, with 6 of the 15 strata including only a single tract group. Possibly as a consequence of this limitation, variation in the estimated prevalence across the 26 tract groups within strata was less than expected by chance, preventing estimation of a clustering effect. We therefore used the 54 census tracts rather than the census tract groups as the PSUs for the primary sampling design. For data analysis we also combined the Young and Old strata among Salt Lake County Low-prevalence Hispanics, and we also combined the Young and Old strata among Salt Lake County Low-prevalence non-Hispanics, due to insufficient census tracts within the individual strata. For the secondary sampling design, we used the more numerous block groups as the PSU in statistical analyses for all strata in which block groups were the true PSUs. For Park City, the household itself was the PSU in the secondary sampling design, and thus the household itself served as the PSU in data analysis.

#### Data Analysis

We constructed Jackknife replicate weights (11), which we then applied in statistical analyses to estimate standard errors and perform statistical inference. The Jackknife approach provides a largely model-free approach for estimating variability while accounting for correlations in outcomes between respondents in the same PSU, and also naturally accounts for the use of different PSUs in the primary and secondary sampling designs. We modeled the relationship of seroprevalence and other outcomes (e.g., self-reported COVID concern, and self-reported social distancing) to predictor variables (e.g., county, demographic and clinical factors, behaviors and attitudes) using survey weighted generalized linear models for a binary outcomes with variability assessed based on the replicate jackknife weights. We implemented these analyses using the Survey package of R. We tested for the presence of a detectable temporal trend in seroprevalance by including calendar time as a continuous variable in models relating seroprevalance to the Utah Department of Health May 20 case count and calendar time. These analyses showed no trend for an effect of calendar time. Hence, analyses for seroprevalance are presented without adjustment for a secular trend in calendar time.

#### Adjusting estimates of seropositivity for assay error

Direct estimates of seroprevalence based on the proportion of tested respondents with positive serology assays are biased because the sensitivity and specificity of the test is expected to be less than 100%. Given relatively low seroprevalence, estimates of seroprevalence are especially strongly affected by the specificity of the test. As recommended by the Abbott ARCHITECT SARS-CoV-2 IgG package insert (4), we estimated specificity as 0.996, based on an evaluation of 1070 samples obtained prior to the COVID-19 outbreak, including 73 samples from individuals with other respiratory illnesses. This evaluation found that the assay incorrectly classified 4 of these 1070 “true negative” samples as positive for COVID-19. We estimated a sensitivity of 0.83 which corresponded to the 25 of 30 respondents who reported having had a positive COVID diagnosis and whose serology results were obtained at least 1 week later and were also positive. In sensitivity analyses we also considered a sensitivity estimate of 0.972, which is the proportion of 107 samples from subjects known to have COVID-19 that led to positive test results (104/107). These 107 samples included 73 from subjects with onset of COVID-19 symptoms at least 14 days prior to the test, and 34 subjects whose onset of COVID-19 symptoms was between 8 and 13 days prior to the test. Given these estimates of sensitivity and specificity, we then provided corrected estimates of seroprevalence by applying the formula: (P1 - (1-specificity))/(sensitivity + specificity - 1), where P1 is the estimated prevalence provided as described above. Finally, we used a parametric bootstrap resampling approach to account for the sampling error in the Abbott estimate of specificity when presenting lower and upper 95% confidence limits.

We did not further expand confidence limits to account for uncertainty in sensitivity. Instead, we conducted sensitivity analyses that evaluated how our estimates of seroprevalences are modified under different assumed values for the true sensitivity, which are compatible with the previous studies described in the above paragraph.

## Figures and Tables

Figure S1 displays the geographic locations of the primary or secondary sampling designs. The figure illustrates that the primary sampling locations are spread across the four counties and that a large fraction of the counties were sampled either in the primary or secondary sampling design.

**Figure S1.**
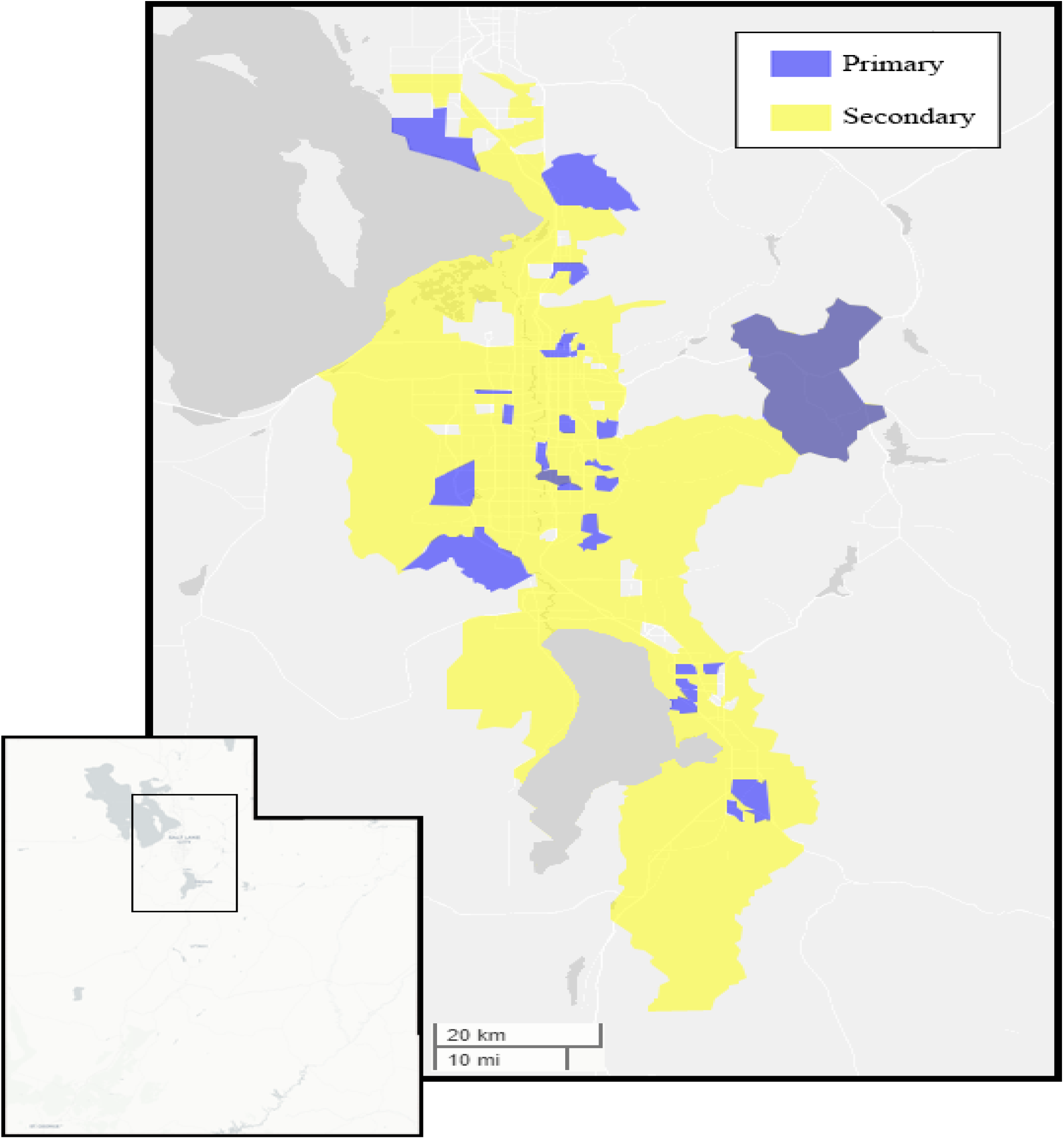
Geographic Areas Sampled in the Primary and Secondary Sampling Designs.

**Figure S2** displays the timing of the serology and PCR samples for the study. The extended sampling design refers to the full collection of the 5,125 responding households, including households in both the primary and secondary sampling designs.

**Figure S2.**
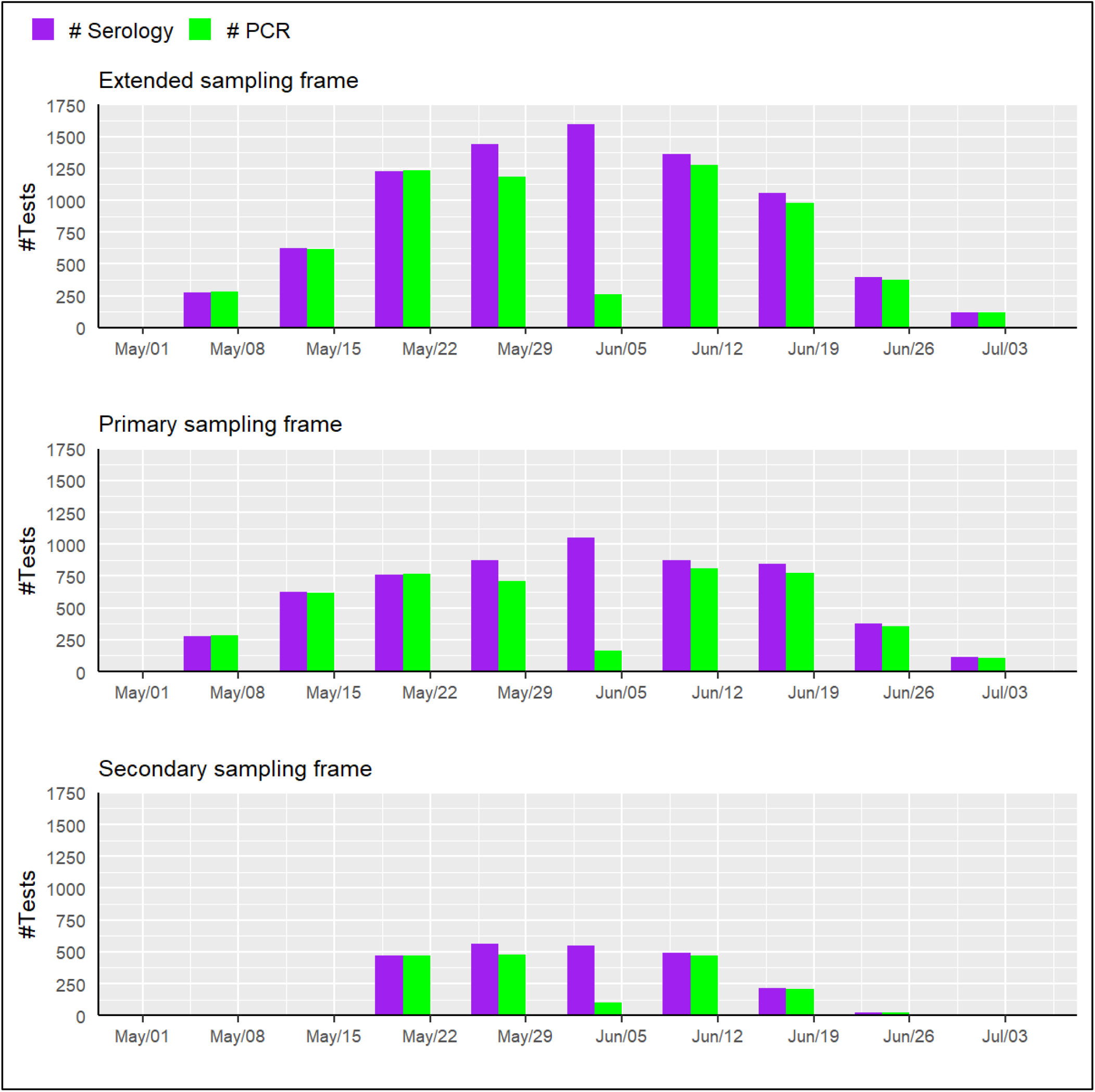
Timing of the Serology and PCR Samples for the Study. **Notes:** The extended sampling design refers to the full collection responding households from both the primary and secondary sampling designs.

**Figure S3** displays the differences in subject characteristics in the primary and secondary sampling designs.

**Figure S3.**
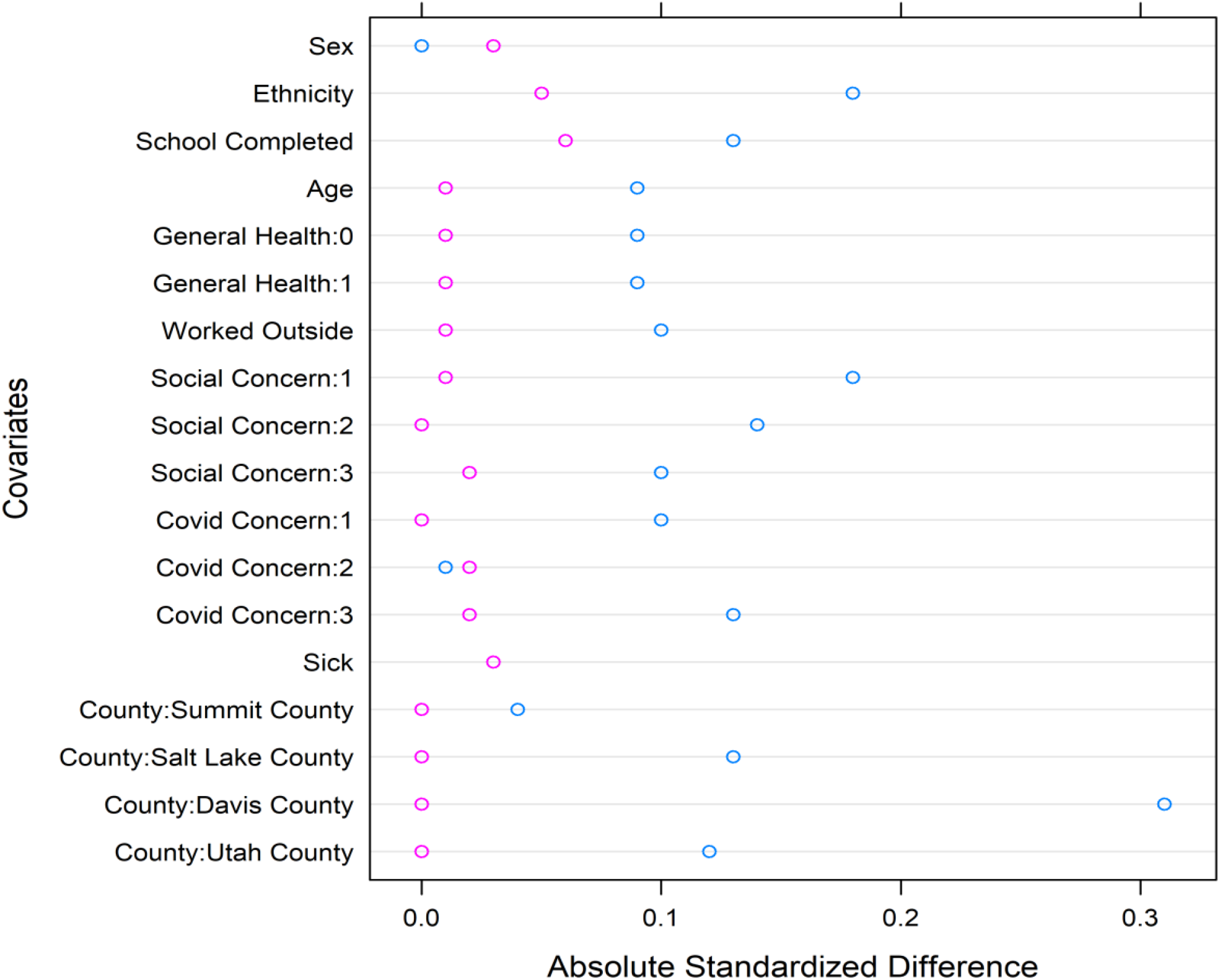
Propensity Matching of Secondary to Primary Sampling Design Respondents. **Notes:** The open blue circles represent the standardized mean differences in each factor between the primary and secondary sampling designs following application of sampling weights that account for nonresponse at the household, individual, and serology testing levels. The open pink circles represent the standardized mean differences after the additional propensity score weighting to bring the characteristics of the respondents in the secondary sampling design into alignment with the characteristics of the respondents in the primary sampling design. The shift in the pink circles relative to the blue circles indicates the impact of the propensity adjustment to align the secondary design sample to match the primary design sample.

**Figure S4** depicts the dependence of the estimated seroprevalence on the sensitivity of the IgG assay. We based our primary estimates of seroprevalence on estimates a sensitivity of 0.83, based on the fraction of 25/30 respondents who self-reported having a prior positive COVID-19 test and subsequently had a positive serology test at least one week subsequent to their reported positive COVID-19 test. We considered a relatively wide range for sensitivity to address speculation that IgG concentrations might wane over time and become undetectable by the assay at some point.

**Figure S4.**
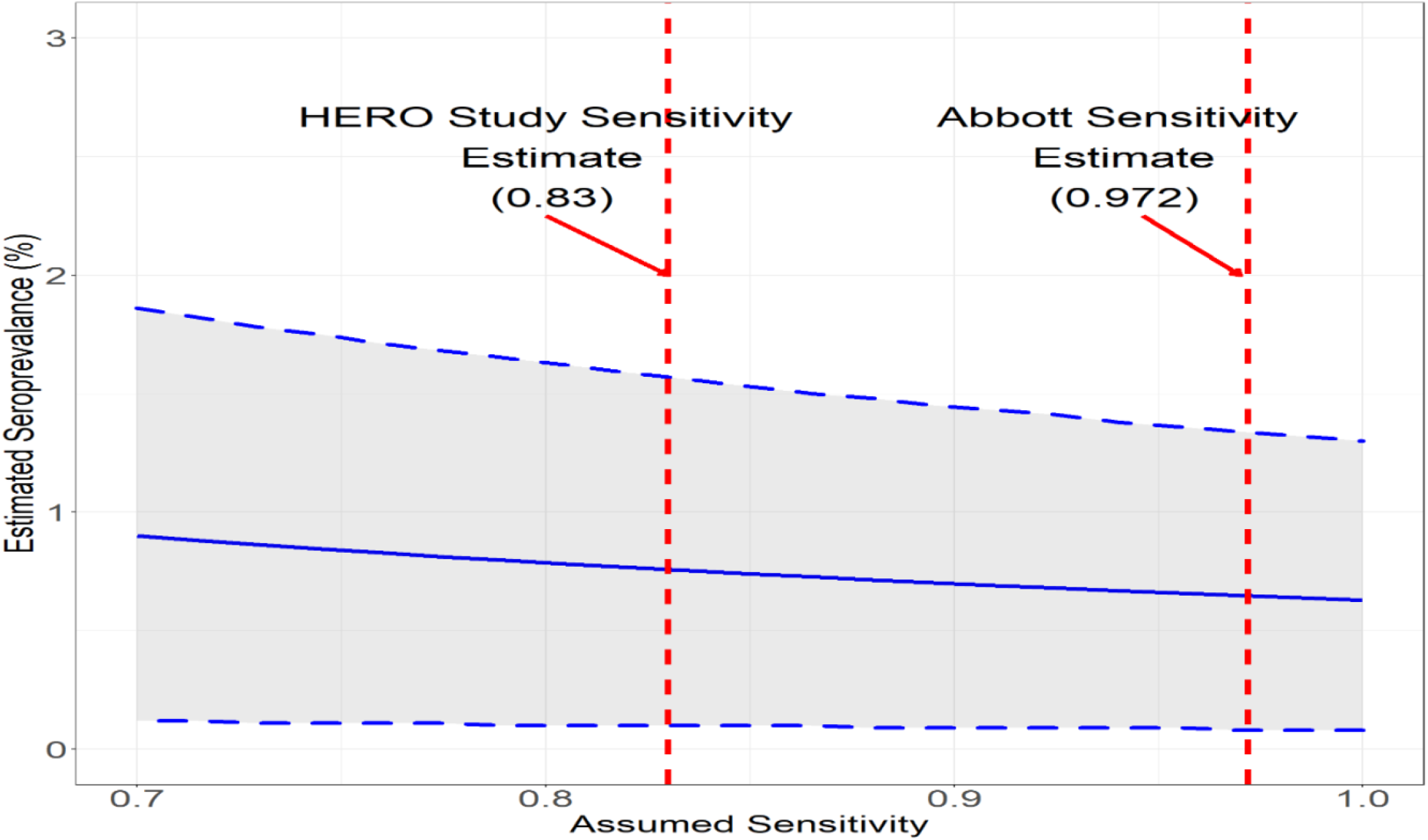
Dependence of Percent Seropositive on Assumed Sensitivity of the Serology Assay for the Analyses. **Notes:** Shown is the relationship between the estimated seroprevalence across the 4-county area with the assumed sensitivity if specificity is assumed to be 0.996.

**Figure S5** displays the incidence of positive PCR tests over the course of the study.

**Figure S5:**
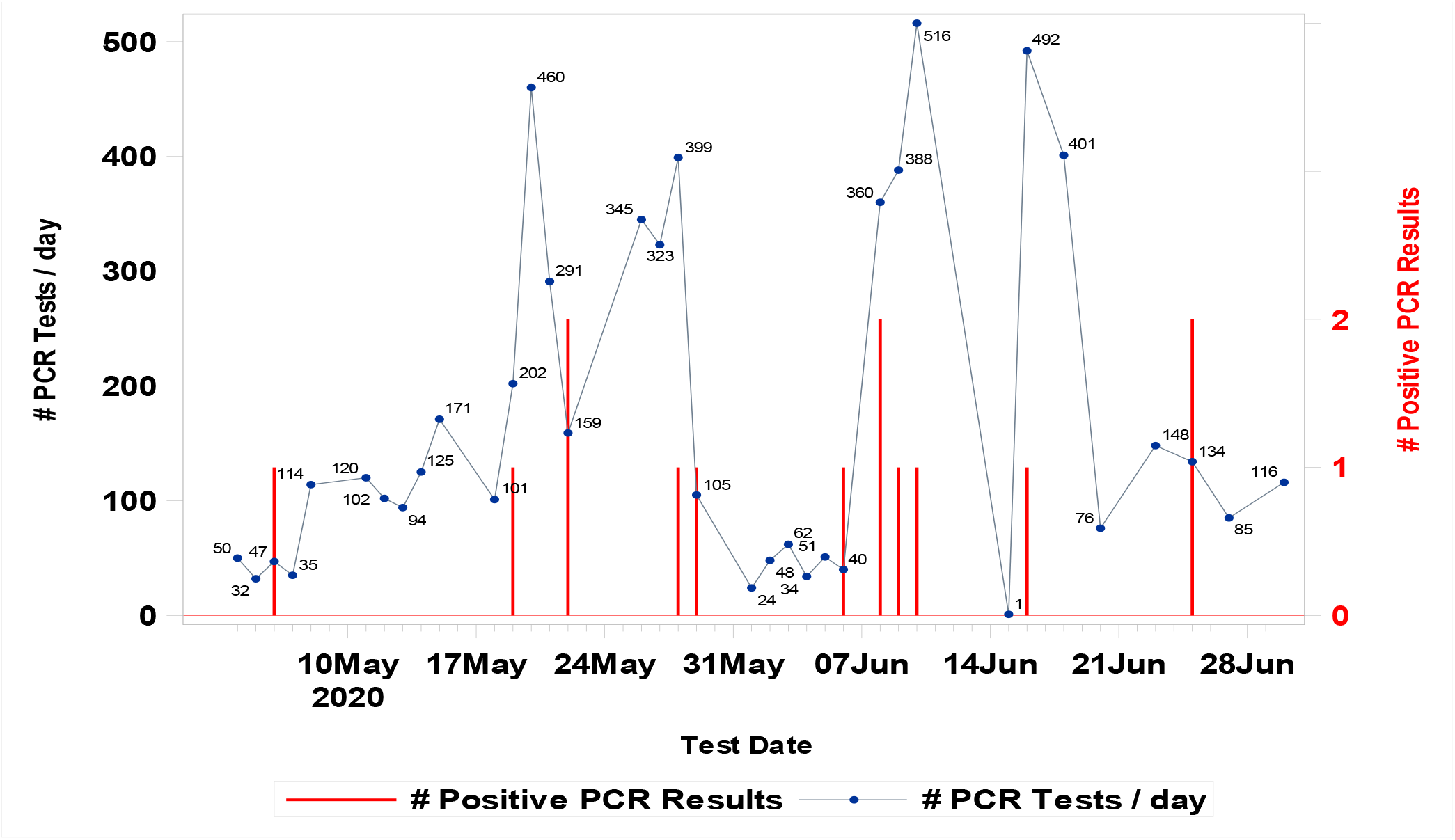
Positive PCR Tests and Total Number of PCR Tests for Study Participants. **Notes:** The grey curve is plotted relative to the left vertical axis and indicates the number of PCR tests performed each day. The drop-off in this curve in late May and early June reflects a temporary period during which PCR tests were administered only if specifically requested by the respondent. The study subsequently reinitiated broad PCR testing in response to the increase COVID-19 case counts reported in the 4-county area. The vertical red bars are plotted relative to the right vertical axis, and indicate the weeks of occurrence of positive PCR tests in the study.

**Table S1.** Predictor Variables in Propensity Score Nonresponse models.

| Household response propensity model | Individual response propensity model | Serology response propensity model |
| --- | --- | --- |
| *Location Predictors*<br>1A) Tract group (primary sampling design only)<br>1B) Serology testing location (secondary sampling design only)<br>*Predictors from US Census*<br>1) % of the population less than or equal to 14 years of age<br>2) Median Age<br>3) % Hispanic<br>4) % not entering college<br>5) % of families with annual income less than \$60,000<br>6) % of families with annual income less than \$40,000 | *Location Predictors*<br>1A) Tract group (primary sampling design only)<br>1B) Serology testing location (secondary sampling design only)<br>*Predictors from US Census*<br>1) Median age,<br>2) % with family income less than \$40,000<br>*Predictors from Household Survey*<br>1) Implements social distancing<br>2) Degree of concern over COVID<br>3) Regularly leaves the home for work, medical treatment, groceries, or to go to restaurants, 4) General health<br>5) Hispanic ethnicity<br>6) Education level<br>7) Has been tested previously for COVID-19<br>8) Degree of concern that others should social distance. | *Location Predictors*<br>1A) Tract group (primary sampling design only)<br>1B) Serology testing location (secondary sampling design only)<br>*Predictors from Individual Survey*<br>1) Implements social distancing<br>2) Degree of concern over COVID<br>3) Regularly leaves the home for work, medical treatment, groceries, or to go to restaurants, 4) General health<br>5) Respondent age<br>6) Respondent sex<br>7) Hispanic ethnicity<br>8) Education level<br>9) Has been tested previously for COVID-19<br>10) Degree of concern that others should social distance. |

**Tables S2-S6** provide response rates for the respective strata in the primary and secondary sampling designs.

**Table S2.** Household Response Rates^*^.

| Stratum | Primary Sampling Design |  |  | Secondary Sampling Design |  |  |
| --- | --- | --- | --- | --- | --- | --- |
|  | No.<br>Responded | No.<br>Approached | %<br>Response | No.<br>Responded | No.<br>Approached | %<br>Response |
| Davis County High-prevalence | 375 | 833 | 45 | . | . | . |
| Davis County Low-prevalence | 374 | 1036 | 36.1 | . | . | . |
| Davis County High/Low-prevalence | . | . | . | 274 | 2125 | 12.9 |
| Salt Lake County High-prevalence<br>Hispanic Old | 364 | 1316 | 27.7 | . | . | . |
| Salt Lake County High-prevalence<br>Hispanic Young | 210 | 834 | 25.2 | . | . | . |
| Salt Lake County High-prevalence<br>Hispanic Young/Old | . | . | . | 186 | 2280 | 8.2 |
| Salt Lake County High-prevalence<br>Non-Hispanic Old | 283 | 868 | 32.6 | 135 | 912 | 14.8 |
| Salt Lake County High-prevalence<br>Non-Hispanic Young | 289 | 876 | 33 | 49 | 462 | 10.6 |
| Salt Lake County Low-prevalence<br>Hispanic Old | 131 | 415 | 31.6 | 36 | 456 | 7.9 |
| Salt Lake County Low-prevalence<br>Hispanic Young | 160 | 412 | 38.8 | 33 | 462 | 7.1 |
| Salt Lake County Low-prevalence<br>Non-Hispanic Old | 471 | 1225 | 38.4 | 146 | 912 | 16 |
| Salt Lake County Low-prevalence<br>Non-Hispanic Young | 157 | 406 | 38.7 | 45 | 462 | 9.7 |
| Summit County | 165 | 876 | 18.8 | 118 | 3205 | 3.7 |
| Utah County High-prevalence<br>Hispanic | 258 | 818 | 31.5 | 88 | 912 | 9.6 |
| Utah County High-prevalence Non-Hispanic | 146 | 416 | 35.1 | 47 | 456 | 10.3 |
| Utah County Low-prevalence<br>Hispanic | 161 | 411 | 39.2 | . | . | . |
| Utah County Low-prevalence Non-Hispanic | 294 | 821 | 35.8 | . | . | . |
| Utah County Low-prevalence<br>Hispanic/Non-Hispanic | . | . | . | 130 | 1368 | 9.5 |

\*In the primary sampling design, we operationally defined household contacts as a visit by the field team or the initiation of the Web Survey in response to the mailer sent to the household. Respondents were households that completed key portions of the household survey or at least one individual survey. We estimated the response rates as the ratio of these two quantities. In the secondary sampling design, we defined household contacts as being sent the mailer. We used different definitions between the two sampling designs because the principal sampling method in the primary sampling design was door-to-door contact by the field team, with mailings playing a secondary role, while in the secondary sampling design the only sampling method was the mailer.

**Table S3.** Individual Response Rates^*^.

| Stratum | Primary Sampling Design |  |  | Secondary Sampling Design |  |  |
| --- | --- | --- | --- | --- | --- | --- |
|  | No. Responded | No. Approached | % Response | No. Responded | No. Approached | % Response |
| Davis County High-prevalence | 741 | 950 | 78 | . | . | . |
| Davis County Low-prevalence | 764 | 1100 | 69.5 | . | . | . |
| Davis County High/Low-prevalence | . | . | . | 576 | 697 | 82.6 |
| Salt Lake County High-prevalence Hispanic Old | 614 | 774 | 79.3 | . | . | . |
| Salt Lake County High-prevalence Hispanic Young | 325 | 505 | 64.4 | . | . | . |
| Salt Lake County High-prevalence Hispanic Young/Old | . | . | . | 348 | 404 | 86.1 |
| Salt Lake County High-prevalence Non-Hispanic Old | 518 | 639 | 81.1 | 275 | 315 | 87.3 |
| Salt Lake County High-prevalence Non-Hispanic Young | 471 | 590 | 79.8 | 96 | 107 | 89.7 |
| Salt Lake County Low-prevalence Hispanic Old | 258 | 340 | 75.9 | 69 | 82 | 84.1 |
| Salt Lake County Low-prevalence Hispanic Young | 314 | 457 | 68.7 | 69 | 83 | 83.1 |
| Salt Lake County Low-prevalence Non-Hispanic Old | 908 | 1233 | 73.6 | 314 | 354 | 88.7 |
| Salt Lake County Low-prevalence Non-Hispanic Young | 340 | 514 | 66.1 | 92 | 99 | 92.9 |
| Summit County | 160 | 177 | 90.4 | 171 | 202 | 84.7 |
| Utah County High-prevalence Hispanic | 524 | 706 | 74.2 | 195 | 234 | 83.3 |
| Utah County High-prevalence Non-Hispanic | 305 | 413 | 73.8 | 124 | 147 | 84.4 |
| Utah County Low-prevalence Hispanic | 352 | 532 | 66.2 | . | . | . |
| Utah County Low-prevalence Non-Hispanic | 598 | 843 | 70.9 | . | . | . |
| Utah County Low-prevalence Hispanic/Non-Hispanic | . | . | . | 312 | 378 | 82.5 |

\*We defined individual response rates in both sampling designs as the proportion individuals age 12 or older in responding households that completed the individual survey.

**Table S4.** Serology Response Rates^*^.

| Stratum | Primary Sampling Design |  |  | Secondary Sampling Design |  |  |
| --- | --- | --- | --- | --- | --- | --- |
|  | No. Responded | No. Approached | % Response | No. Responded | No. Approached | % Response |
| Davis County High-prevalence | 593 | 746 | 79.5 | . | . | . |
| Davis County Low-prevalence | 594 | 791 | 75.1 | . | . | . |
| Davis County High/Low-prevalence | . | . | . | 516 | 598 | 86.3 |
| Salt Lake County High-prevalence Hispanic Old | 512 | 651 | 78.6 | . | . | . |
| Salt Lake County High-prevalence Hispanic Young | 201 | 348 | 57.8 | . | . | . |
| Salt Lake County High-prevalence Hispanic Young/Old | . | . | . | 287 | 361 | 79.5 |
| Salt Lake County High-prevalence Non-Hispanic Old | 429 | 534 | 80.3 | 245 | 289 | 84.8 |
| Salt Lake County High-prevalence Non-Hispanic Young | 352 | 489 | 72 | 87 | 100 | 87 |
| Salt Lake County Low-prevalence Hispanic Old | 217 | 272 | 79.8 | 63 | 69 | 91.3 |
| Salt Lake County Low-prevalence Hispanic Young | 227 | 332 | 68.4 | 60 | 73 | 82.2 |
| Salt Lake County Low-prevalence Non-Hispanic Old | 732 | 958 | 76.4 | 274 | 320 | 85.6 |
| Salt Lake County Low-prevalence Non-Hispanic Young | 252 | 356 | 70.8 | 83 | 93 | 89.2 |
| Summit County | 218 | 277 | 78.7 | 127 | 179 | 70.9 |
| Utah County High-prevalence Hispanic | 441 | 554 | 79.6 | 171 | 195 | 87.7 |
| Utah County High-prevalence Non-Hispanic | 261 | 329 | 79.3 | 124 | 141 | 87.9 |
| Utah County Low-prevalence Hispanic | 288 | 363 | 79.3 | . | . | . |
| Utah County Low-prevalence Non-Hispanic | 474 | 619 | 76.6 | . | . | . |
| Utah County Low-prevalence Hispanic/Non-Hispanic | . | . | . | 280 | 331 | 84.6 |

\*We defined serology response rates in both sampling designs as the proportion of individual survey respondents who also provided a serology sample.

**Table S5.**
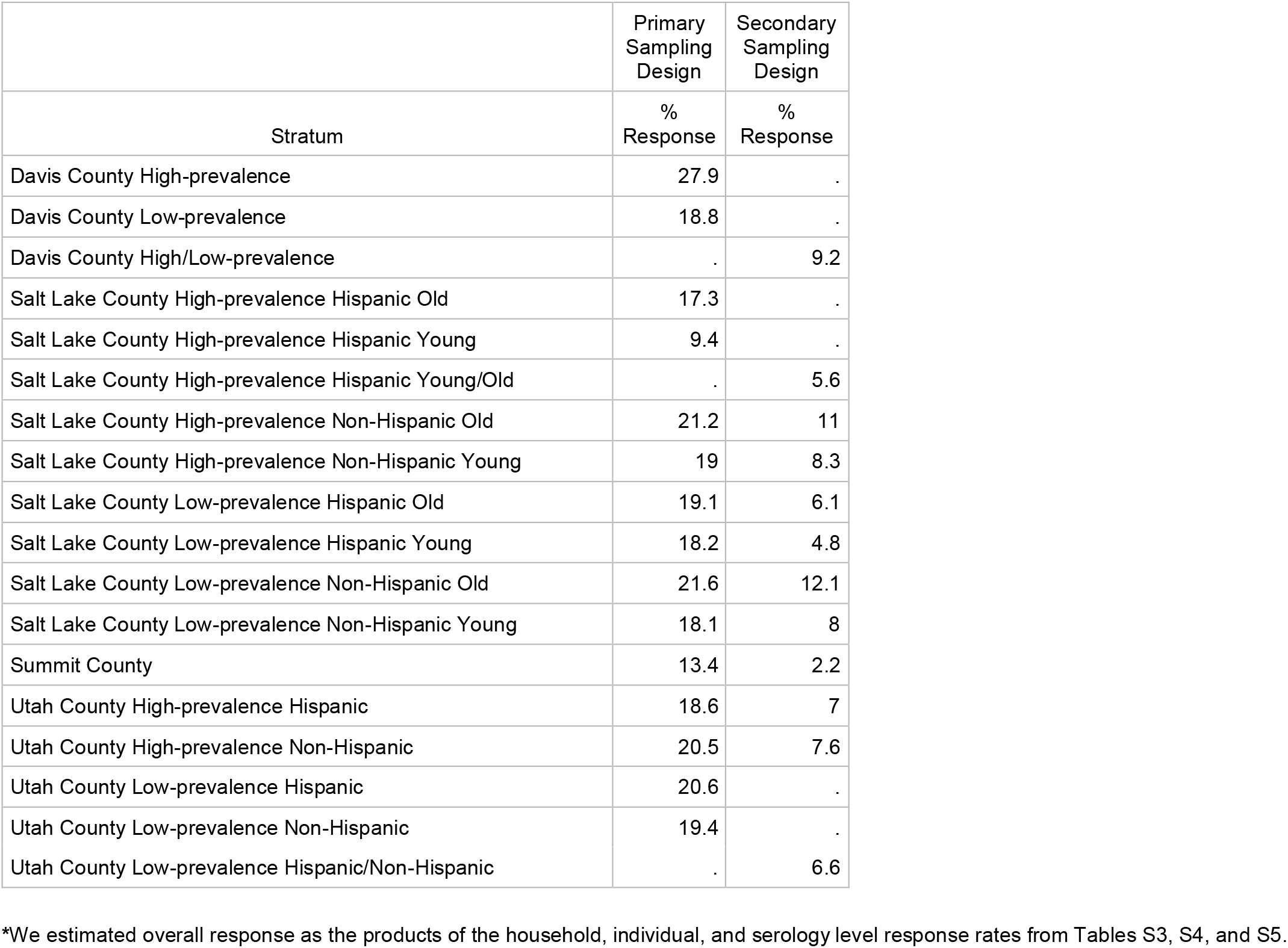
Overall Response Rates^*^.

|  | Primary<br>Sampling<br>Design | Secondary<br>Sampling<br>Design |
| --- | --- | --- |
| Stratum | %<br>Response | %<br>Response |
| Davis County High-prevalence | 27.9 | . |
| Davis County Low-prevalence | 18.8 | . |
| Davis County High/Low-prevalence | . | 9.2 |
| Salt Lake County High-prevalence Hispanic Old | 17.3 | . |
| Salt Lake County High-prevalence Hispanic Young | 9.4 | . |
| Salt Lake County High-prevalence Hispanic Young/Old | . | 5.6 |
| Salt Lake County High-prevalence Non-Hispanic Old | 21.2 | 11 |
| Salt Lake County High-prevalence Non-Hispanic Young | 19 | 8.3 |
| Salt Lake County Low-prevalence Hispanic Old | 19.1 | 6.1 |
| Salt Lake County Low-prevalence Hispanic Young | 18.2 | 4.8 |
| Salt Lake County Low-prevalence Non-Hispanic Old | 21.6 | 12.1 |
| Salt Lake County Low-prevalence Non-Hispanic Young | 18.1 | 8 |
| Summit County | 13.4 | 2.2 |
| Utah County High-prevalence Hispanic | 18.6 | 7 |
| Utah County High-prevalence Non-Hispanic | 20.5 | 7.6 |
| Utah County Low-prevalence Hispanic | 20.6 | . |
| Utah County Low-prevalence Non-Hispanic | 19.4 | . |
| Utah County Low-prevalence Hispanic/Non-Hispanic | . | 6.6 |

\*We estimated overall response as the products of the household, individual, and serology level response rates from Tables S3, S4, and S5.

**Table S6** summaries the mean relative weights applied to various subgroups of respondents in our final analyses. These weights incorporate adjustments for nonresponse at the household, individual, and serology levels, followed by the propensity score adjustment to align the characteristics of respondents to the secondary sampling design to the primary sampling, as well as the final iterative proportional fitting step to alight the weighted characteristics of the study population to the US census.

**Table S6.**
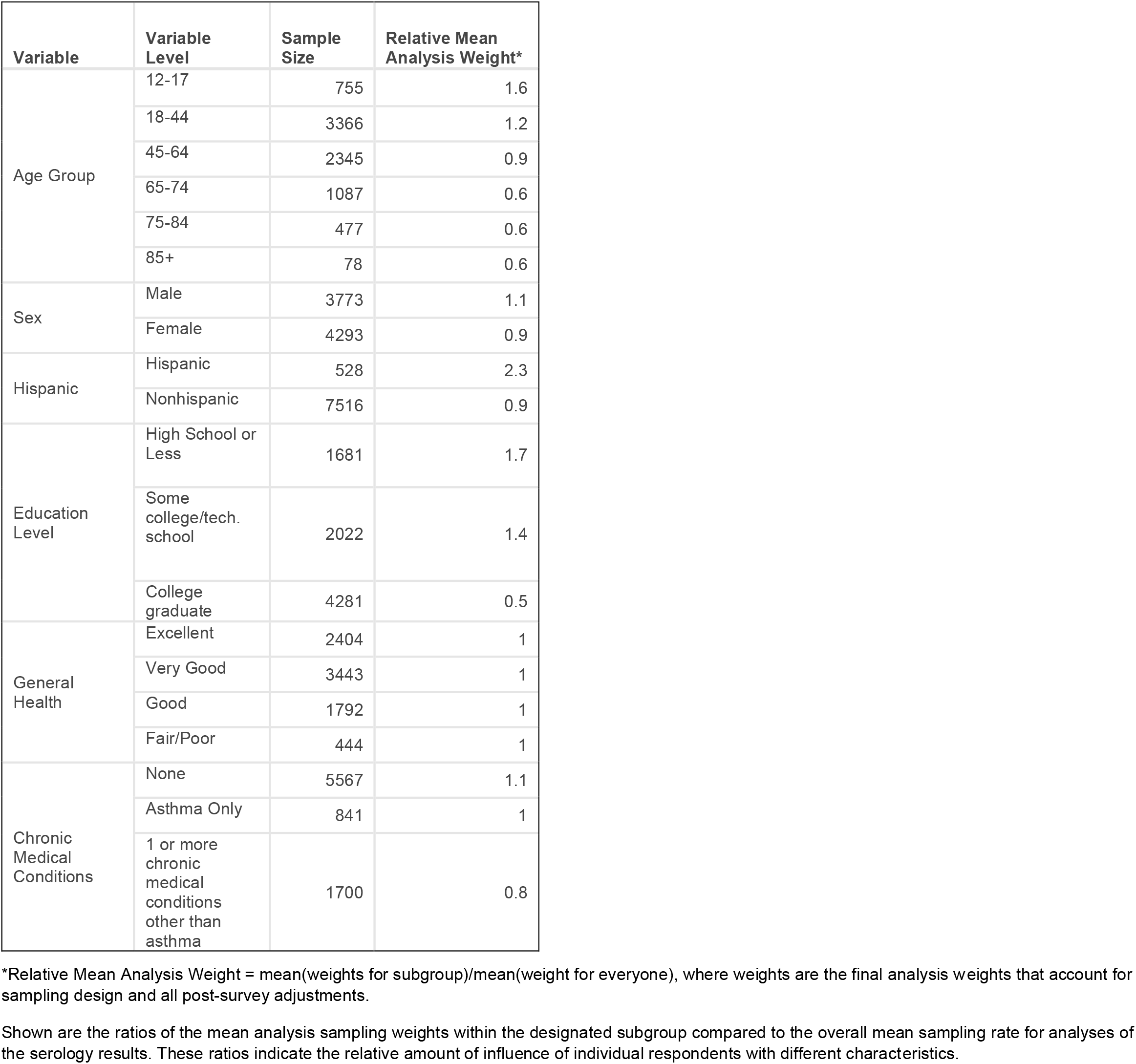
Analysis Weights for Serology Analyses by Respondent Characteristics.

